# Circulating levels of leptin in different phases of the menstrual cycle – a systematic review and meta-analysis

**DOI:** 10.1101/2023.08.28.23294657

**Authors:** Suhrud J. Panchawagh, Pratyush Kumar, Shabarini Srikumar, Khushi Prajapati, Manali Sarkar, Kumar Abhishek, Poonam Agrawal, G. P. Kaushal, Daniel Jesús García Martínez, Swati Kalra, Euden Bhutia, Aman Agarwal, Kunika Singh, Shubhangi Sharma, Tejaswini Ashok, Urmil Shah, Rushikesh Shukla, Sejal Gupta, Hareem Shaikh, K. Sri Soumya, Poorvikha S

**Affiliations:** Smt. Kashibai Navale Medical College & General Hospital, Pune; ReQuir Statistics Solutions; Dr Baba Saheb Ambedkar Medical College and Hospital, Delhi, India; Tirunelveli Medical College and Hospital, Tirunelveli; Hinduhridayasamrat Balasaheb Thackeray Medical College & Dr.R.N.Cooper Municipal General Hospital, Mumbai; Francisco De Vitoria University, Madrid, Spain; Maulana Azad Medical College, New Delhi; Xinjiang Medical university, China; BJ Government Medical College and Sassoon General Hospitals, Pune; JSS Medical College, Mysore; Rajiv Gandhi Medical College and Chhatrapati Shivaji Maharaj, Thane; Jawaharlal Nehru Medical College, Datta Meghe Institute of Medical Science, Swangi, Wardha-442001, Maharashtra, India; JIIU’S Indian institute of medical sciences and research, Jalna; SDM College of Medical Sciences and Hospital, Dharwad; St John’s Medical College, Bangalore

**Keywords:** Leptin, menstrual cycle, follicular phase, ovulatory phase, luteal phase

## Abstract

Leptin is a peptide hormone synthesized by white adipose tissue. Its principal site of action is in the brainstem and hypothalamus, where it acts to modulate satiety and control of reward and aversion. Leptin deficiency or resistance is associated with dysregulation of cytokine production, increased susceptibility to infections, autoimmune disorders, malnutrition, and inflammatory responses. Studies have shown variation in leptin levels between males and females, and in different phases of the menstrual cycle as well. Our study aims to explain the variation in leptin levels and their physiological role in the menstrual cycle. A total of 1887 records were identified from PubMed, Google Scholar, and Cochrane. We included studies where females were 18-40 years of age, who had achieved menarche, and were on regular menstrual cycles. We excluded studies which were cross-section, pre-prints, or studies where inadequate information was provided. Seven studies were included in the study. We used the standardized mean difference as the outcome measure. Leptin levels varied significantly in the phases of the menstrual cycle as well as during prepubertal, pubertal, and postpubertal phases. The standardized mean difference in leptin levels between the follicular phase and the luteal phase was 2.67 ng/mL (95% CI: 0.16 to 5.19 ng/mL). Fluctuations in leptin levels can cause weight changes and reproductive cycle disturbances. Hence, in cases of infertility with abnormal menstrual cycles, measurement of the physiologic variability in leptin levels may help to diagnose the cause of infertility and appropriate interventions can be started.

## Background

The LEP or ob/obesity gene (located on chromosome 7q31.3) synthesizes Leptin, which is a peptide hormone.^[1]^ Yellow adipose tissue is the primary site for secretion of leptin in humans and other mammals.^[2]^ Leptin receptors (LepRs) have various isoforms, which are expressed in several body tissues, including the hypothalamus, choroid plexus, ovary, uterus, testis, lung, kidney, liver, adrenal gland, placenta, peripheral blood mononuclear cells, and the white and brown adipose tissues.^[3]^ The role of leptin in regulation of energy homeostasis, neuroendocrine function, and metabolism is established. As it is an adipocytokine, the concentration of leptin in the circulation is directly proportional to adipose tissue mass; consequently, it conveys information about the available body energy stores to the brain.^[3]^ A previous Scottish study has demonstrated that the mean serum level of leptin is two to three times higher in women than men with the same body fat mass.^[4]^ A relation in concentrations of circulating fertility hormones and circulating leptin in female subjects of reproductive age can explain the gender difference.^[5]^ In addition, some studies propose that alterations in body fat and circulating levels of leptin contribute in initiating the onset of puberty.^[6]^ A study by Elias C.F further suggest that increased concentrations of leptin might be one of the factors, which contribute towards the trend of decrease in age of onset of puberty in obese girls.^[7]^ On the other hand, the levels of leptin in postmenopausal women are found to be almost equal to that of men of same age and body fat percentage.^[8]^ Hence, it has become evident that leptin is strongly associated with fertility and reproductive functions in females. Moreover, a change in concentration of leptin in different phases of the menstrual cycle has been observed in many studies. There are, however, reports that suggest that there is no significant change in circulating leptin levels with respect to estradiol and progesterone in the different phases.^[9]^

Leptin was first discovered in 1994, since then multiple studies were undertaken to understand the role of leptin in obesity and related disorders.^[10,11]^ The idea of involvement of leptin in female fertility came between 1998 and 1999, when it was discovered that leptin levels were varying significantly in women at different periods of life, and were the highest during the reproductive period.^[10]^ Leptin has also been tried as a modality of treatment for hypothalamic infertility.^[7]^ Most studies on leptin levels in different phases of the menstrual cycle have shown leptin to have the highest concentration during the luteal phase.^[12,13]^ While some studies suggest that it also rises significantly just prior to ovulation.^[13]^ The relation between leptin and other fertility hormones has however been unclear to previous researchers^[9,13,14]^

Our main aim is to identify the cause of variation in circulating levels of leptin during different phases of the menstrual cycle and to check for heterogeneity of results among different studies. In addition, we aim to condense and synthesize explanations for why leptin levels differ in different phases of the cycle, and possible explanations for the physiological role of leptin in different phases of the menstrual cycle, to pave the way for understanding menstrual cycle pathologies better.

## Objectives

- Our primary objective was to assess whether serum leptin levels change during various phases of the menstrual cycle.
- Our secondary objectives were to assess whether other serum gynecological hormones change during various phases of the menstrual cycle.

## Methods

### Criteria for considering studies for this review

Type of studies: Randomized or quasi-randomized controlled trials of changes in serum leptin levels in different phases of the menstrual cycle were to be included. Unpublished data and abstracts were eligible for inclusion provided adequate information regarding primary outcomes could be obtained from them.

### Types of participants

All females aged 18-40 years who achieved menarche and were on regular menstrual cycles (that repeats every 21-40 days) were included.

### Types of interventions

Measurement of serum leptin levels at least in the follicular and luteal phases in women in the trials.

### Types of outcome measures

Primary outcomes:

1. Serum leptin levels in the follicular and luteal phases of the menstrual cycle (ng/mL)

Secondary outcomes:

1. Serum leptin levels in the follicular, ovulatory, and luteal levels of the menstrual cycle.
2. Serum vaspin levels in different phases of the menstrual cycle.
3. Serum adiponectin levels in different phases of the menstrual cycle.

### Search methods for identification of studies

Electronic searches: PubMed, Cochrane Central Register of Controlled Trials (CENTRAL), and Google Scholar were searched using the following combinations of keywords using Boolean operators:

> “Leptin” AND “Menstrual cycle” OR
>
> “Receptors, Leptin” AND “Menstrual cycle” OR
>
> “Leptin levels” AND “Menstrual cycle” OR
>
> “Leptin” AND “Menstrual cycle phases” OR
>
> “Receptors, Leptin” AND “Menstrual cycle phases” OR
>
> “Leptin levels” AND “Menstrual cycle phases” OR
>
> “Leptin levels” AND “Menstrual cycle phases” AND “Body Mass Index” OR
>
> “Vaspin” AND “Menstrual cycle” OR
>
> “Vaspin” AND “Menstrual cycle phases” OR
>
> “Adiponectin” AND “Menstrual cycle” OR
>
> “Adiponectin” AND “Menstrual cycle phases”

We used a PICOT approach for searching the database. Using these keywords, we searched the databases mentioned. Only English databases were searched as mentioned in the inclusion criteria. All the included studies from the databases were added to the screening software Rayyan.ai.

Reports and studies were screened and reviewed by 4 reviewers independently working under one group. All the reports were reviewed manually by the reviewers, no automated selection tools were used. A total of 7 studies made it to final screening. Full texts were available for all 7 included studies.

One reviewer independently collected data from the reports. No automation tool or software was used to extract data. All the reports had some inconsistencies regarding the duration of menstrual phases considered, population age group, and BMI of the population. Certain inclusion criteria (BMI <26 and Age 18-40 years) were used to uniformly include the data from the reports, and a standardized duration of phases was used.

Outcome domains that were used in our review were the mean Leptin Concentration (ng/ml), Phase of menstrual cycle at which the Leptin was measured, Age, and BMI of the population. The outcome domains were modified in the range to fit the results of maximum studies. Some studies that could not provide the outcome domains were excluded from the review. The mean Leptin concentration, Phase of the menstrual cycle, and BMI were the most important outcome domains to reach conclusions in our study.

All studies that talked about leptin concentration and correlated it with phases of the menstrual cycle were taken into consideration.

The study also considered the BMI, Age, and Regularity of the Menstrual cycle of subjects included. We didn’t need the necessary correlation with other hormones.

Papers were searched and reviewed manually by 3 researchers that worked as a part of a team project. No automated tools were used.

Data was sought into variables named author name, mean leptin concentration, standard deviation, phase of the menstrual cycle, and sample size during each phase. Certain studies which did not mention the duration of certain menstrual phases were assumed according to the standard duration of that phase of the menstrual cycle.

Meta-analysis of all the variables was done using R software ver. 4.2.2 using the ‘metafor’ statistical package.^[15]^

Risk of bias assessment: Cochrane RoB assessment tool version 2 was used for risk assessment. One reviewer used the risk assessment to assess the risk of bias. No other automation tool was used.

### Inclusion criteria

We included a wider range of studies to better answer our research question. All randomized controlled trials, cross-sectional studies, and prospective cohort studies were included. Peer-reviewed studies published in English which reported the leptin concentration in a comprehensive format were included. The study population of regularly menstruating healthy females in the reproductive age group was included. The study group BMI was <26, age 18-40 years, non-breastfeeding women for the last 6 months, not pregnant, and without any gynecological disorder.

### Exclusion criteria

Unpublished manuscripts, abstracts, and conference abstracts were not included. Studies that did not report the leptin concentration phase-wise were excluded. Females suffering from irregular menstrual periods, or gynecological disorder, were excluded. Populations with BMI >26 were also excluded.

### Data collection and analysis

Selection of studies: All published articles identified as potentially relevant by the literature search were assessed for inclusion in the review by the search team of 4 authors. Retrieved articles were assessed and data were abstracted. Discrepancies regarding inclusion and exclusion of the studies were resolved by consensus.

### Data extraction and management

For included studies, data regarding study methodology (including the method of randomization, blinding, drug intervention, stratification, and whether the trial was single or multicenter) and information regarding trial participants (including age groups of females, and other inclusion and exclusion criteria) was extracted.

### Assessment of risk of bias in included studies

The methodological quality of each identified trials was assessed with respect to: 1) Randomization process, 2) Deviations from the intended interventions, 3) Missing outcome data, 4) Measurement of the outcome, 5) Selection of the reported result

### Statistical analysis and measures of effect for change in leptin levels

We used R software ver. 4.2.2 for MS Windows. The analysis was carried out using the standardized mean difference as the effect measure. A random-effects model was fitted to the data. The amount of heterogeneity (i.e., tau²), was estimated using the restricted maximum-likelihood estimator. In addition to the estimate of tau², the Q-test for heterogeneity and the I² statistic are reported (I2 < 25 %: weak heterogeneity; I2 = 25-50 %: moderate heterogeneity; I2 > 50 %: large or extreme heterogeneity). In case any amount of heterogeneity is detected (i.e., tau² > 0, regardless of the results of the Q-test), a prediction interval for the true outcomes is also provided. Studentized residuals and Cook’s distances are used to examine whether studies may be outliers and/or influential in the context of the model. Studies with a studentized residual larger than the 100 × (1 - 0.05/(2 X 7))th percentile (=99.64%) of a standard normal distribution are considered potential outliers (i.e., using a Bonferroni correction with two-sided alpha = 0.05 for 7 studies included in the meta-analysis). Studies with a Cook’s distance larger than the median plus six times the interquartile range of the Cook’s distances are considered to be influential. The rank correlation test and the regression test, using the standard error of the observed outcomes as predictor, are used to check for funnel plot asymmetry.

## Results

The initial literature search yielded a total of 1887 studies in accordance with the topic. A total of 667 records were eliminated in the screening. The remaining potential studies were evaluated as per a systematic literature search strategy using the preferred reporting items for systematic reviews and meta-analysis (PRISMA) guidelines. With the application of exclusion and inclusion criteria, we have been able to narrow down 7 studies. (Figure 1)

**Figure 1:**
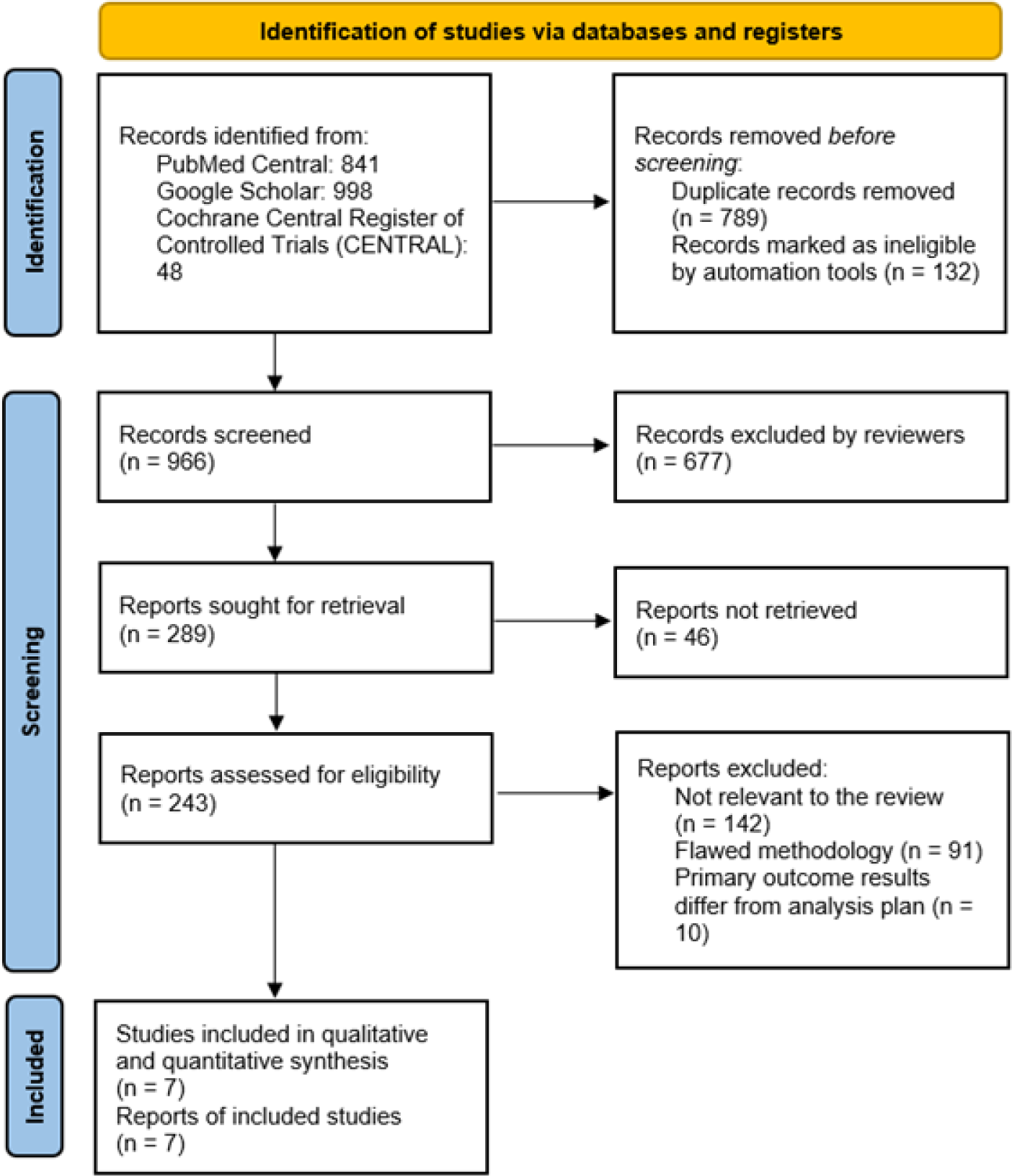
PRISMA flow-chart for the evaluation of the studies.

Qualitative synthesis of the 7 studies included in our systematic review and meta-analysis was performed. The results are provided in Table 1.

**Table 1:**
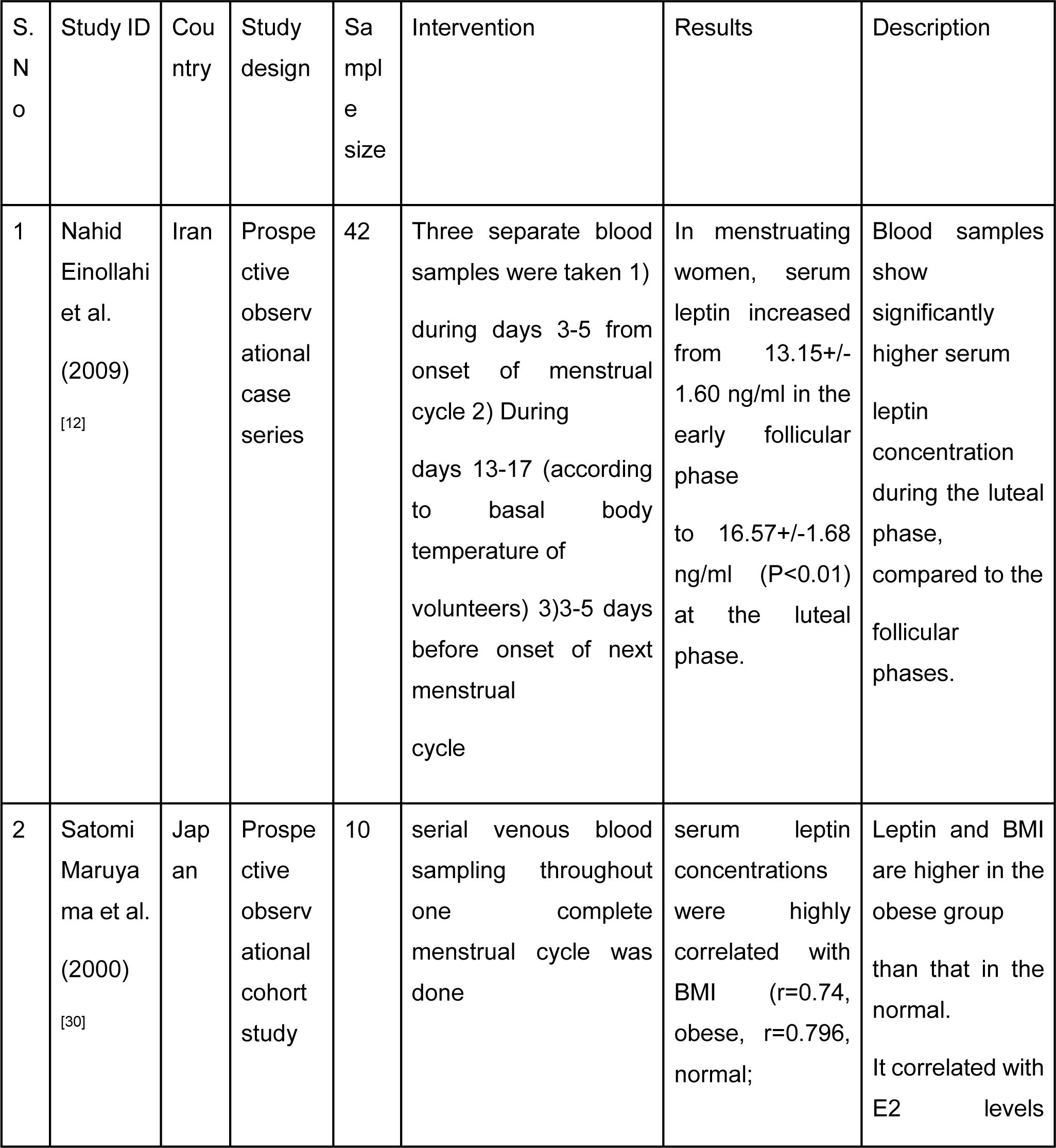
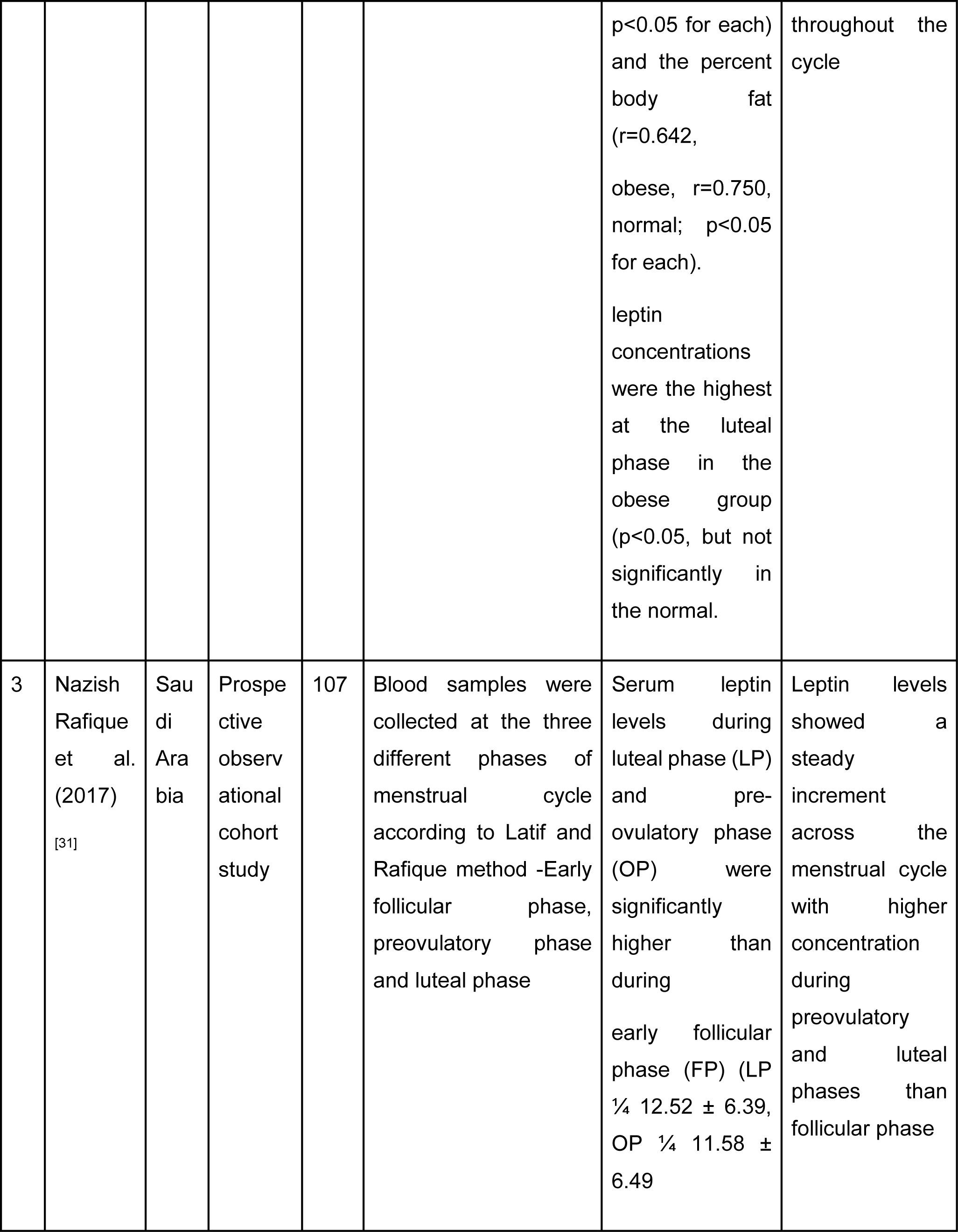
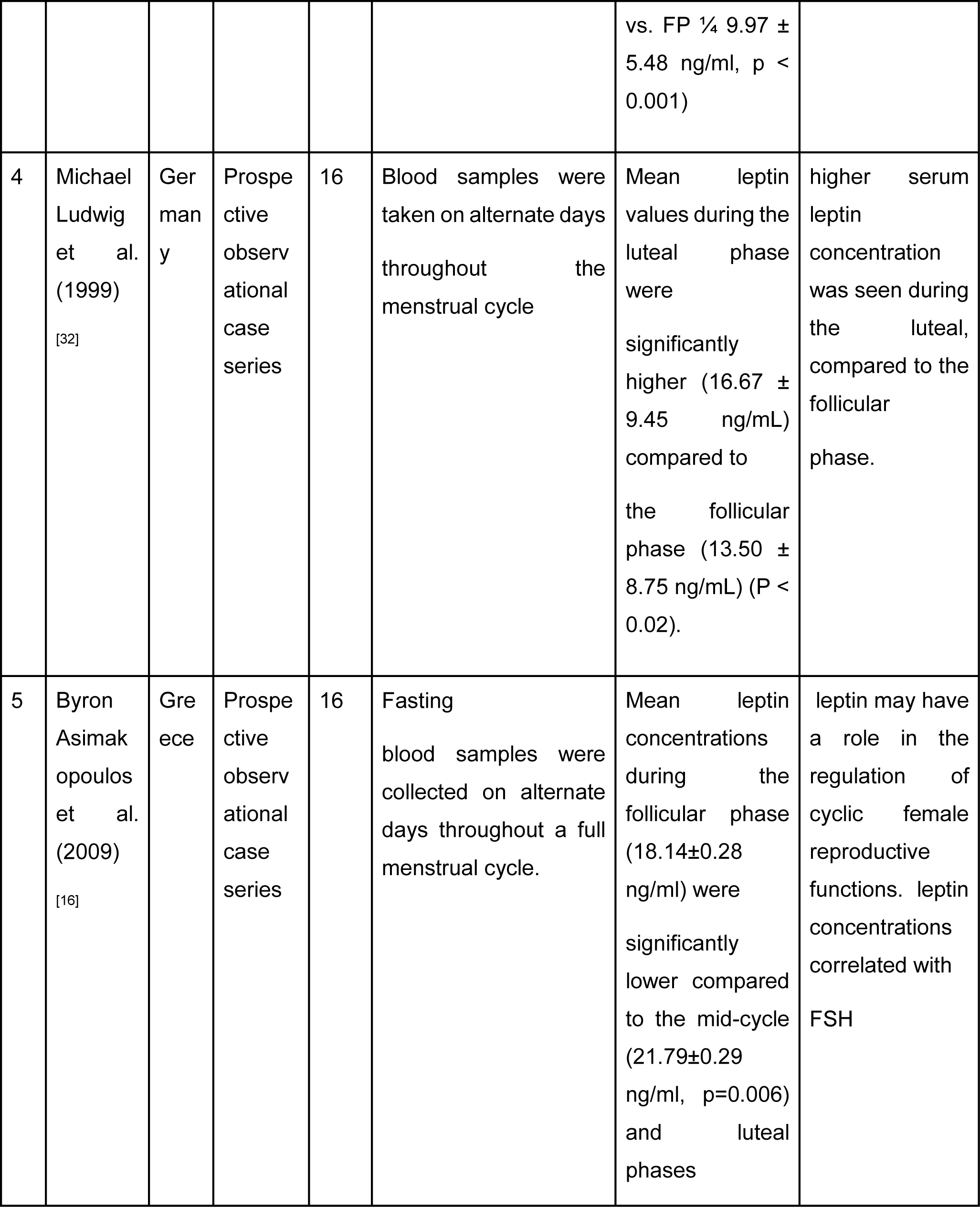
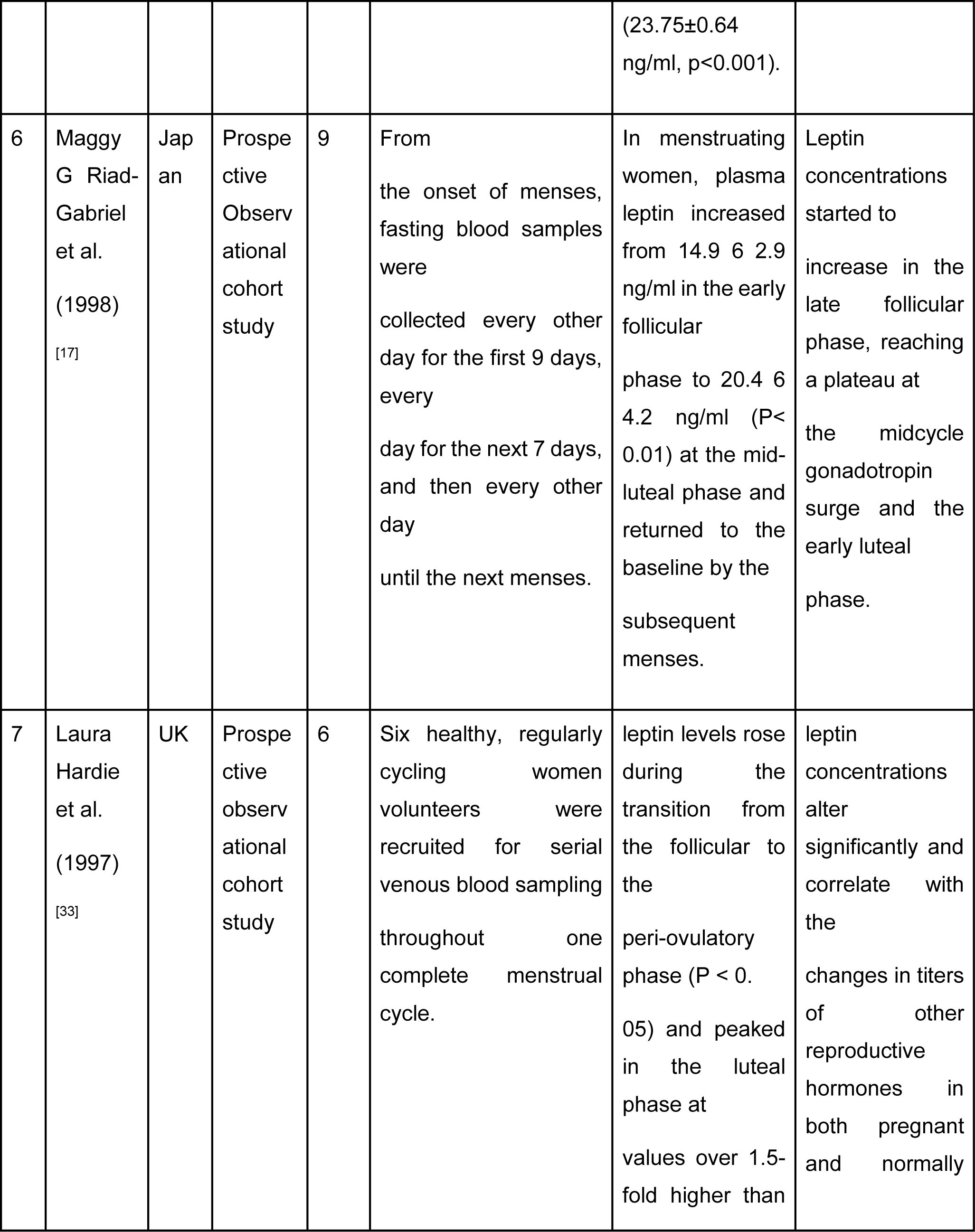
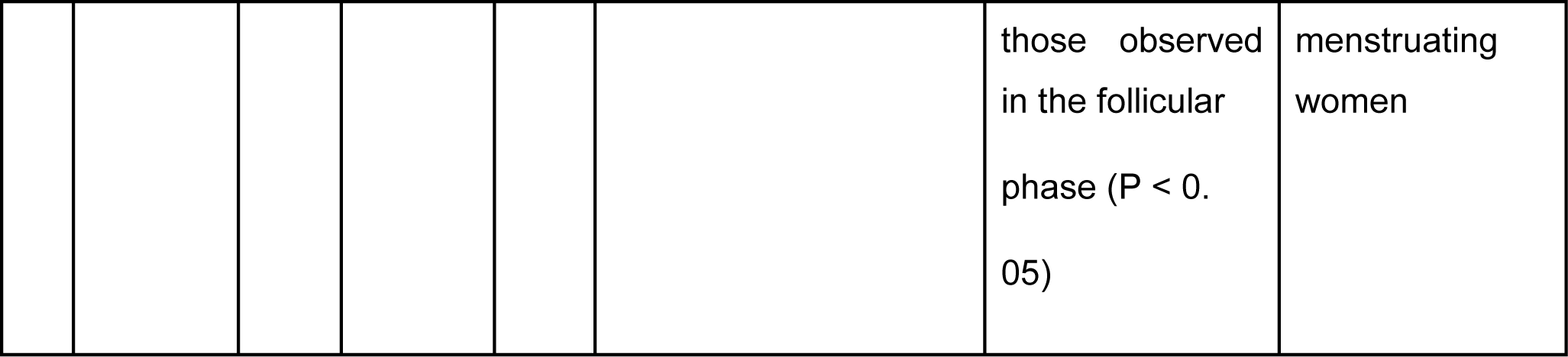
Qualitative synthesis of results. This table consists of 7 studies which fit in our inclusion criteria. The details about the study design, sample size, intervention, results, and description have been mentioned in the table below.

### Meta-analysis

A total of k=7 studies were included in the analysis. The observed standardized mean (of leptin levels [in ng/mL]) differences ranged from -11.0704 to -0.4219, with the majority of estimates being negative (100%). The estimated average standardized mean difference based on the random-effects model was -2.6729 (95% CI: -5.1890 to -0.1569). (Figure 2) Therefore, the average outcome differed significantly from zero (z = -2.0822, p = 0.0373). According to the Q-test, the true outcomes appear to be heterogeneous (Q(6) = 71.8255, p < 0.0001, tau² = 11.0075, I² = 98.2746%). A 95% prediction interval for the true outcomes is given by -9.6454 to 4.2995. Hence, although the average leptin levels in the follicular phase is estimated to be lesser than the luteal phase, in some studies the true outcome may in fact be greater. An examination of the studied residuals revealed that one study (Byron Asimakopoulos et al.) had a value larger than ± 2.6901 and may be a potential outlier in the context of this model. According to Cook’s distances, one study (Byron Asimakopoulos *et al*.) could be considered to be overly influential.^[16]^ The regression test indicated funnel plot asymmetry (p < 0.0001) but not the rank correlation test (p = 0.2389). (Figure 3) Results of the risk of bias assessment as performed by Cochrane’s Risk of Bias Assessment Tool version 2 are provided in Figure 4.

**Figure 2:**
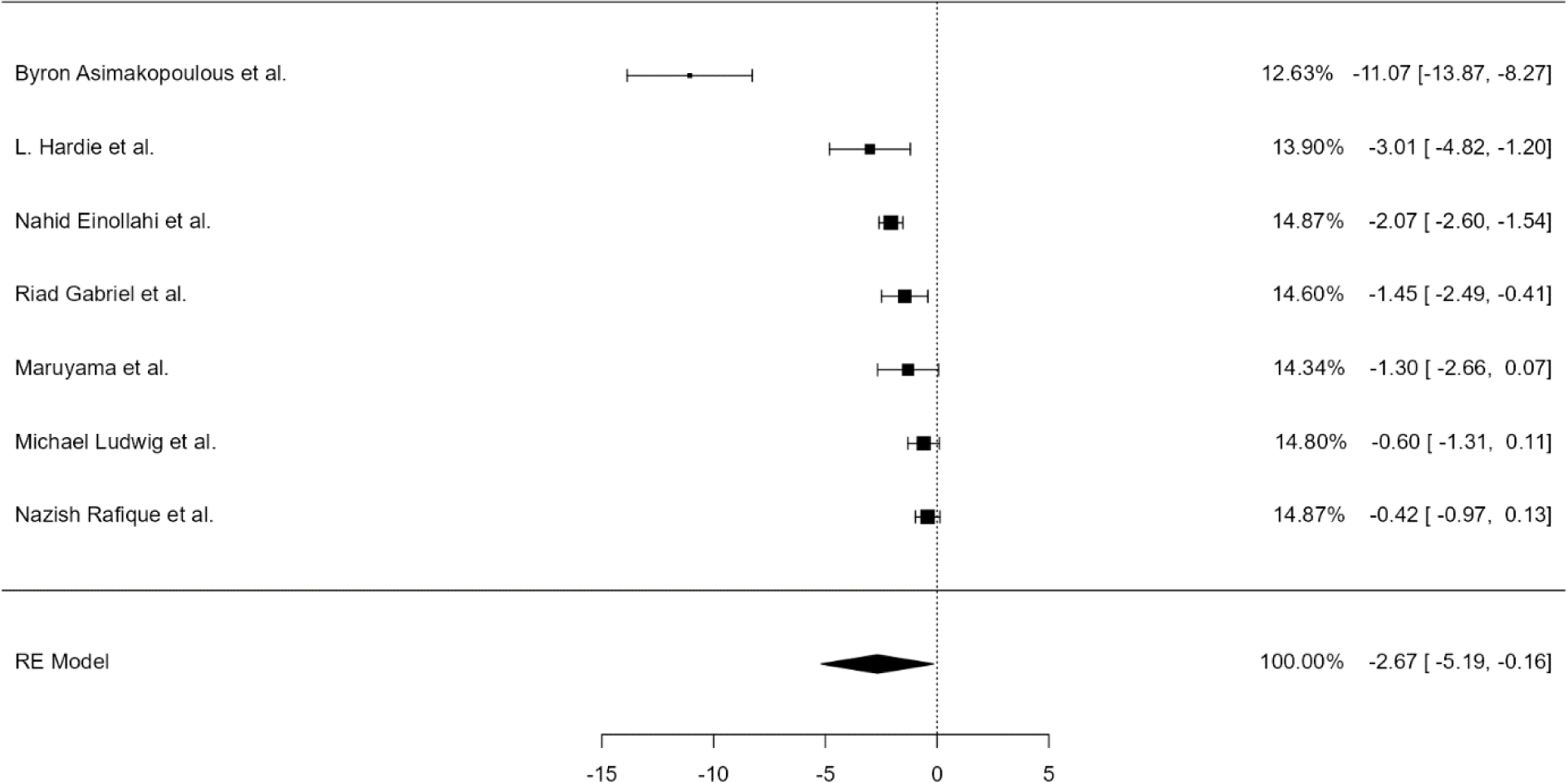
Forest plot of change in serum leptin levels from the follicular phase to the luteal phase. Positive values indicate higher leptin levels in the follicular phase than the luteal phase, while negative values indicate higher leptin levels in the luteal phase than the follicular phase

**Figure 3:**
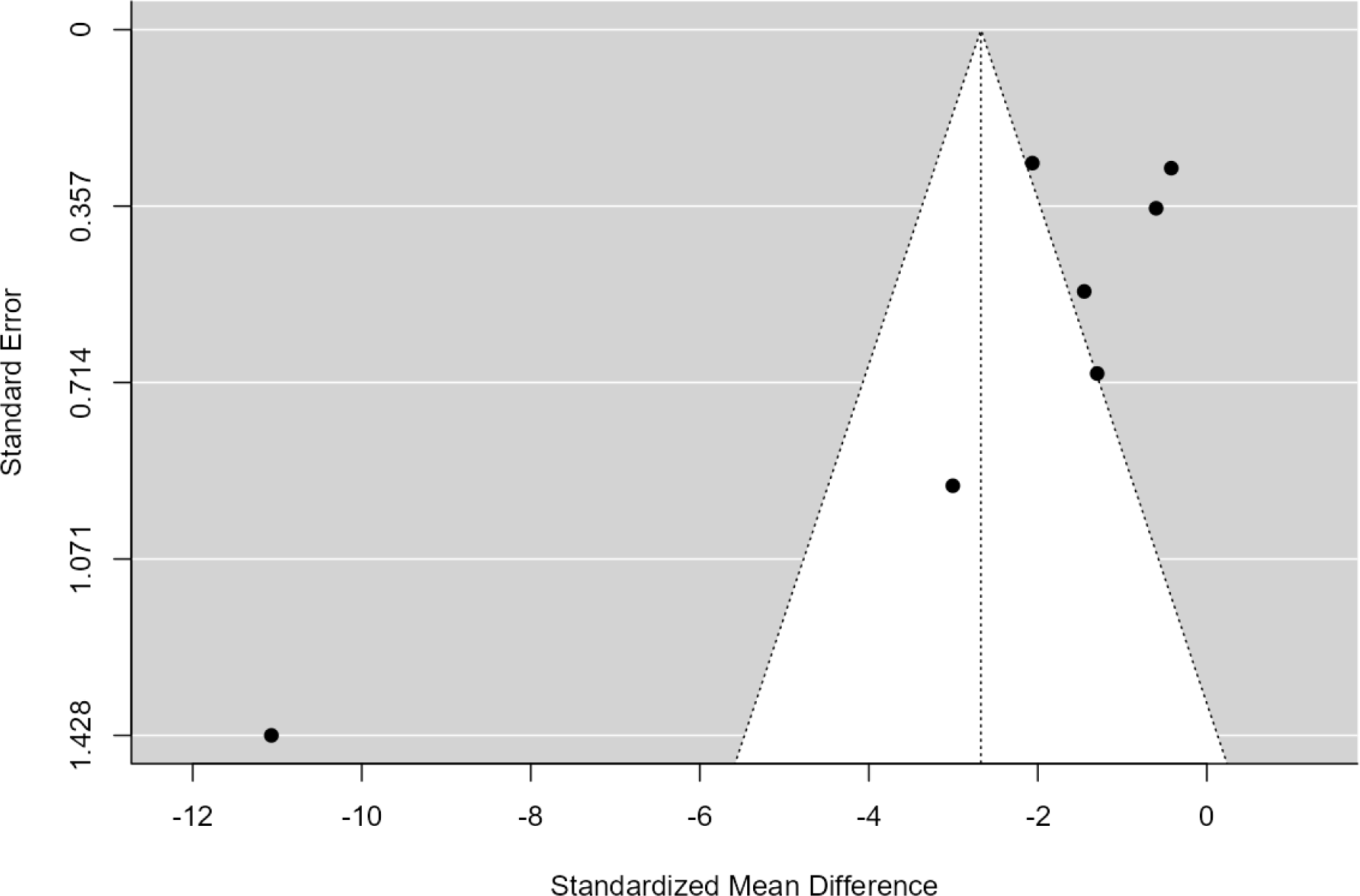
Funnel plot of the standardized mean difference on the X-axis plotted against its standard error on the Y-axis, to assess publication bias.

**Figure 4:**
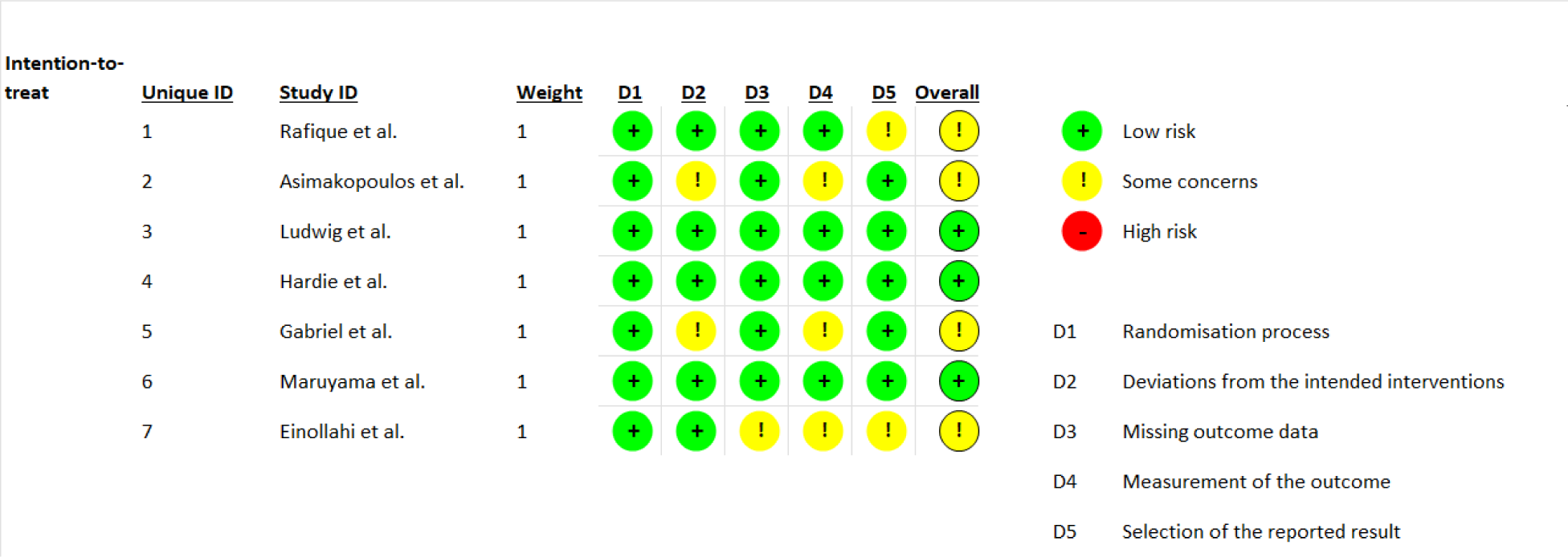
Assessment of the risk of bias using Cochrane’s risk of bias assessment tool 2.

## Discussion

Leptin levels vary significantly in every phase of the menstrual cycle, as well as even during prepubertal and pubertal and post-pubertal periods. Serum leptin level is also affected by various transitional phases of reproduction too; it has been seen that leptin concentration is nearly three-fold higher in post-pubertal girls than in pre-pubertal girls. Leptin concentrations during puberty appear to be partly due to a stimulatory effect of estradiol.^[17]^ Moreover, there is evidence to conclude that serum leptin level was at the lowest level during the menstrual phase and the highest level was around the luteal phase.^[8]^ Positive correlations were found between mean values of leptin and mean values of estradiol during the second half of the follicular phase; this can suggest that estradiol may be involved in the regulation of leptin production in women.^[11]^ Leptin levels are considered more stable on cycle days 1-5 than later in the cycle. A recent study demonstrated that, leptin levels varies during the course of spontaneous cycles in women, elevated during pregnancy, whereas a modest elevation of the same is noted during IVF treatment, possible weight gain from week 13-32 during pregnancy can be correlated with the rise in leptin levels, this weight gain showcases relationship between body mass index and circulating leptin, the best correlation occurring during the luteal phase when progesterone and leptin concentrations are highest.^[13]^ In postmenopausal women, serum leptin levels were positively associated with bone mineral density and bone mineral content, overall, high serum leptin levels were associated with higher BMD levels.^[14]^ Certain studies have however demonstrated that postmenopausal women who are receiving hormone replacement therapy (HRT) have had reduced serum leptin levels in contrast with the woman without HRT who have increased serum leptin level possibly as a consequence of the increase in fat mass.^[18]^

Obesity impacts reproductive functions in both men as well as women. Serum leptin levels are highly correlated with body mass index (BMI) and percent body fat. These levels are also higher in the obese group than in the normal group, due to which serum leptin levels are significantly higher in obese individuals than non-obese ones. This condition had been labeled hyperleptinemia. In eating disorders such as bulimia nervosa, it has been concluded that there is a significant decrease in leptin levels.^[19]^ Similarly recent reports show that congenital leptin deficiency leads to hyperphagia and excessive weight gain from early infancy as well as failure of pubertal onset in adolescence.^[20,21]^ In patients with acute anorexia nervosa, it has been recorded that serum leptin levels were lower than in age-matched controls. A previous hypothesis suggests that amenorrhea in anorexia nervosa is related to the low leptin secretion that has been discussed.^[22]^ Leptin, is produced mainly adipocytes, is elevated in individuals with obesity and may mediate the association between obesity and pregnancy outcomes, it has been seen that women with higher baseline preconception leptin levels had a higher likelihood of experiencing some adverse pregnancy outcomes including gestational diabetes mellitus and hypertensive disorders of pregnancy.^[23]^ However, changes in serum leptin levels of pregnant women with type 1 diabetes mellitus are associated with parallel changes in maternal body weight and their glycemic control, according to the studies, women with low blood pressure had the lowest serum leptin levels throughout pregnancy.^[24]^

In pregnant women, estradiol and leptin are found to have a strong positive correlation.^[25]^ The alterations in serum leptin during the phases of menstrual cycle, however, have not been significantly associated with alterations in serum estradiol and progesterone.^[26]^ Some studies have suggested a positive correlation between concentration of leptin and estradiol during the follicular phase, but there is not enough evidence to confirm the findings.^[27]^

Fluctuations in leptin levels can cause weight changes and reproductive cycle disturbances. Both, deficiency of, and resistance to leptin can lead to infertility. A study on role of leptin in unexplained infertility in north Indian women by Pratibha Kumari *et. a*l showed that regulating leptin concentrations can in fact, control infertility that is attributed to obesity.^[28]^ Resistance to leptin causes increase in concentration of circulating leptin which further explains why women with infertility of unexplained etiology higher circulating concentrations of leptin when compared to fertile women. This relationship between leptin and infertility can be attributed to leptin’s action on the hypothalamic pituitary axis. It can further be noted that lower concentrations of leptin in circulation, such that after rapid weight loss or prolonged fasting, and anorexia nervosa is associated with amenorrhea. Moreover, administration of leptin to infertile women has shown to restore normal menstrual cycle and ovarian function.^[23]^

Leptin administration has also proved to be beneficial in case of congenital and acquired insulin resistance.^[24]^ It acts both centrally and peripherally and it has established its role, not only in treating hyperinsulinemia, but also hypercholesterolemia and hypertriglyceridemia. The association between leptin and insulin resistance also supports the relation between leptin and fertility as insulin resistance is a well-known factor causing impaired fertility in both females and males.^[29]^

## Data Availability

PubMed, Cochrane Central Register of Controlled Trials (CENTRAL), and Google Scholar

## Authors’ conclusions

Women had higher levels of leptin during the preovulatory and luteal phases. Increase in BMI leads to reduced leptin levels. Fluctuations in leptin levels can cause weight changes and reproductive cycle disturbances. In cases of infertility with abnormal menstrual cycles, measurement of the physiologic variability in leptin levels may help to diagnose the cause of infertility and appropriate interventions can be started.

